# Insulin-AKT1-YBX1 Regulation of ANGPTL8 Promote Lipogenesis in Obstructive Sleep Apnea-Associated Dyslipidemia

**DOI:** 10.1101/2024.06.23.24309370

**Authors:** Yuenan Liu, Haolin Yuan, Anzhao Wang, Shengming Wang, Xu Xu, Junhui Hu, Jinhong Shen, Yiming Hu, Xinyi Li, Niannian Li, Zhenfei Gao, Xiaoxu Zhang, Xiaoman Zhang, Yupu Liu, Huajun Xu, Hongliang Yi, Jian Guan, Zhiqiang Li, Yongxu Zhao, Shankai Yin, Feng Liu

## Abstract

Dyslipidemia is a hallmark of obstructive sleep apnea (OSA)-induced metabolic syndrome, yet the mechanisms remain poorly understood. We conducted a genome-wide association study on lipid traits in the OSA cohorts, identifying the SNP rs3745683 in *ANGPTL8*, significantly associated with reductions in multiple lipid traits. ANGPTL8, an essential lipogenic hormone and potential therapeutic target for metabolic syndrome, showed elevated expression in OSA patients compared to healthy controls, strongly correlated with increased insulin levels. Notably, *ANGPTL8* expression can be upregulated by insulin stimulation, indicating it as an insulin-responsive hormone regulating dyslipidemia in OSA. Mechanistically, SNP rs3745683 attenuated *ANGPTL8* transcription by inhibiting its binding to transcription factor YBX1. Insulin prompted AKT1 to phosphorylate YBX1 at Ser102, facilitating YBX1’s nuclear translocation and subsequent regulation of *ANGPTL8* expression and lipid synthesis. Specific knockdown of YBX1 in mouse liver confirmed its necessity for *ANGPTL8* expression and hepatic lipid synthesis in vivo. Our findings highlight *ANGPTL8* as a critical regulator of dyslipidemia in OSA patients, offering a promising therapeutic avenue for managing metabolic syndrome in OSA.

## Synopsis

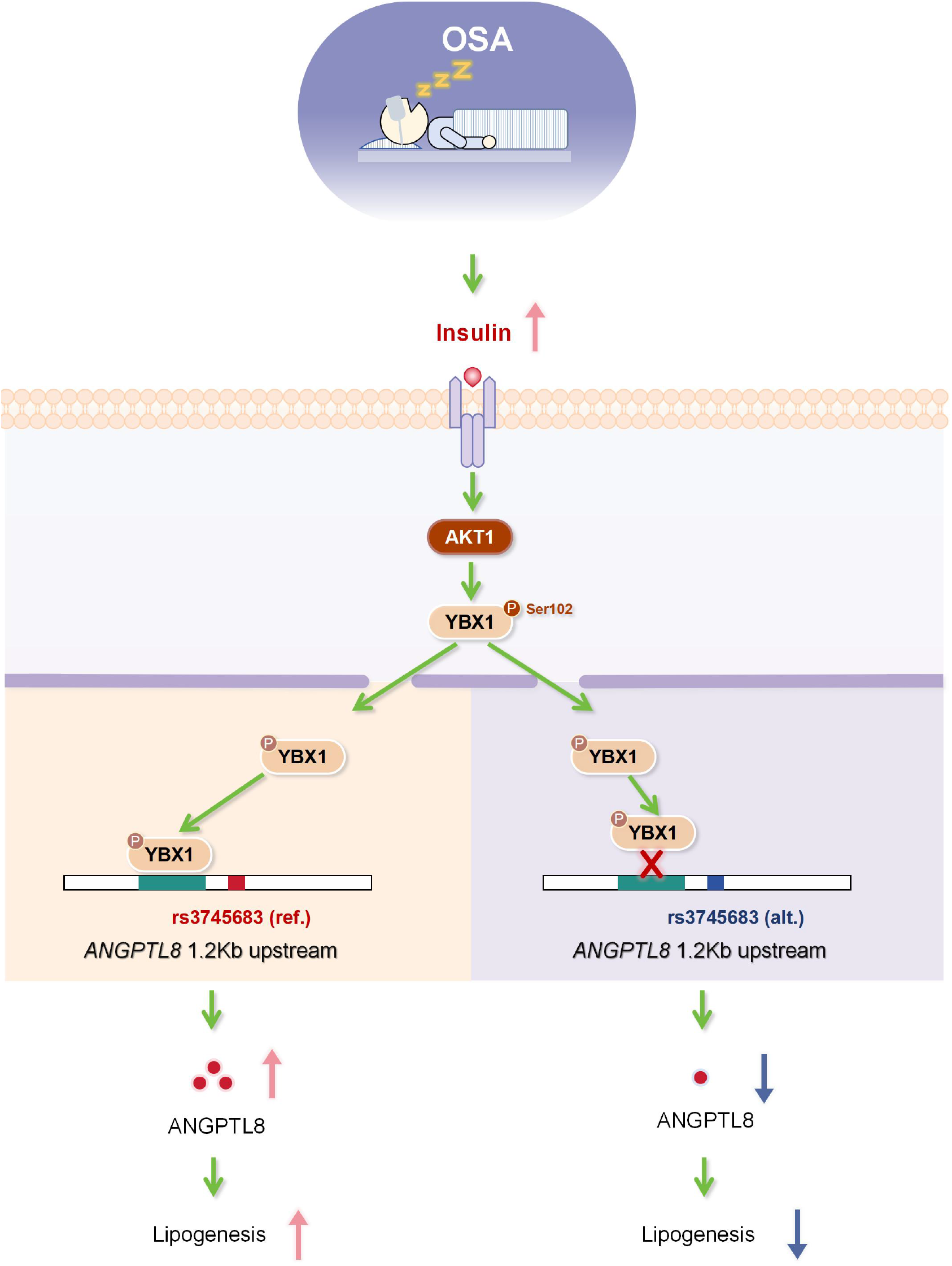

ANGPTL8 serves as a crucial regulator of lipid metabolism abnormalities in obstructive sleep apnea. OSA elevates insulin levels, which, through the AKT1-YBX1 signaling pathway, interact with genetic variants in OSA patients to influence the expression of ANGPTL8 and lipid synthesis.

◆ The genetic variant rs3745683 in ANGPTL8 is associated with lipid profiles in patients with obstructive sleep apnea (OSA).
◆ Elevated insulin levels observed in OSA patients enhance the expression of ANGPTL8 and lipid synthesis.
◆ Insulin signaling facilitates the phosphorylation and nuclear translocation of YBX1, which interacts with rs3745683, collectively influencing the expression of ANGPTL8 and lipid biosynthesis.

## The paper explained

### Problem

Obstructive sleep apnea (OSA) increases the risk of dyslipidemia, which can lead to serious cardiovascular complications. However, the underlying mechanisms remain unclear.

### Results

The GWAS identified a single nucleotide polymorphism (SNP), rs3745683, in ANGPTL8, which is significantly associated with decreases in multiple lipid traits in the OSA cohort. Deficiency of ANGPTL8 led to impaired lipogenesis in hepatocytes and adipocytes. Rs3745683 attenuated ANGPTL8 transcription by suppressing the binding of its proximal element to the upstream transcription factor YBX1. OSA patients exhibited elevated insulin levels, which were highly correlated with increased ANGPTL8 expression. In response to insulin, AKT1 interacted with YBX1 and phosphorylated it at Ser102, promoting its nuclear translocation and subsequent regulation of ANGPTL8 expression and lipid synthesis. Specific knockdown of YBX1 in mouse liver confirmed that YBX1 is required for ANGPTL8 expression and hepatic lipid synthesis in vivo.

### Impact

Our findings highlight ANGPTL8 as a critical regulator of dyslipidemia in OSA patients. We demonstrated that the insulin-AKT1-YBX1 signaling pathway, in conjunction with genetic variations, modulates ANGPTL8 expression and lipid metabolism in OSA-associated dyslipidemia and offered a promising therapeutic avenue for managing metabolic syndrome in OSA patients.

## Introduction

Metabolic syndrome (MS) is distinguished by the co-occurrence of numerous metabolic abnormalities, including central obesity, hyperglycemia (comprising impaired glucose tolerance and diabetes), hyperinsulinemia, hypertension, and dyslipidemia. It constitutes a significant predisposition to cardiovascular disease (CVD) [1]. Substantial research consistently underscores that individuals afflicted with obstructive sleep apnea (OSA), a prevalent sleep-disordered breathing disorder characterized by chronic and intermittent hypoxia as well as enduring sleep deprivation, exhibit a notably heightened susceptibility to developing MS when juxtaposed with the general populace [2–4]. Furthermore, there is compelling evidence that OSA considerably augments the risk of manifesting a spectrum of comorbidities associated with metabolic syndrome, while concurrently contributing to escalated all-cause mortality [3, 5]. Nonetheless, the exact molecular underpinnings governing these relationships remain elusive and necessitate further scholarly inquiry.

Alterations in endocrine hormones represent pivotal factors in the genesis of metabolic syndrome. Angiopoietin like 8 (ANGPTL8), a hepatic and adipose tissue-secreted protein hormone with evolutionary conservation across multiple species, plays an indispensable role in metabolic regulation [6]. A growing body of research, founded on clinical data and murine models, substantiates the central role of ANGPTL8 as a responsive agent in the domains of food intake, estrogen, and insulin signaling within the intricate terrain of glycemic and lipid metabolism [7, 8]. This multifaceted protein exhibits close associations with a spectrum of human conditions, including dyslipidemia [9], obesity [10], metabolic syndrome [10, 11], cardiovascular maladies [12], type 2 diabetes mellitus (T2DM) [13], and obstructive sleep apnea [14, 15]. In its current sphere of investigation, ANGPTL8, operating as an endocrine hormone, orchestrates a symphony of effects by modulating the physiological functions of diverse organs and tissues throughout the body, including the liver [16, 17], pancreas [18], muscles [19], and adipose tissue [19, 20]. Collectively, these studies contribute to the intricate tapestry of metabolic processes that oversee glycemic and lipid regulation.

In the human context, the regulation of metabolism and the etiology of metabolic diseases frequently bear a discernible genetic signature [21–24]. Comprehensive human genetic inquiries, encompassing whole-genome association analyses, irrefutably substantiate the intimate interconnection between *ANGPTL8* and a gamut of metabolic disorders [25–28]. Moreover, these investigations unveil a multitude of genetic loci within *ANGPTL8* intricately intertwined with aberrations in lipid metabolism [25–28]. Among these loci, rs2278426 (C>T), a non-synonymous single nucleotide polymorphism (SNP) within *ANGPTL8,* correlates with decreased serum levels of low-density lipoprotein cholesterol (LDL-C) and high-density lipoprotein cholesterol (HDL-C) in African Americans and Hispanics [29]. Another SNP, rs3745683 (G>A), situated in the *ANGPTL8* promoter region, has been associated with serum HDL-C levels [26]. However, past investigations have provided limited insights into how genetic variations in *ANGPTL8* interact with physiological or pathological factors, thereby elucidating the specific mechanisms by which it modulates the biological functions governing glucose and lipid metabolism.

In our antecedent study, leveraging the Shanghai Sleep Health Study (SSHS) cohort, comprising 5,438 PSG-confirmed OSA patients and 15,152 non-OSA healthy controls, we conducted an extensive genome-wide association study (GWAS). Within this ambit, we identified 18 significant loci linked to susceptibility to OSA and OSA-related quantitative traits, encompassing parameters such as sleep architecture, respiratory events, and oxygen saturation [30]. Concurrently, within the purview of this research undertaking, we conducted a GWAS analysis on the metabolic profiles of the same cohort. This analysis unveiled a notable correlation between a SNP rs3745683(G>A) in the *ANGPTL8* promoter region and a diverse array of metabolic traits, including but not limited to HDL-C, TC, and ApoA. In delving further into the transcriptional regulatory mechanisms governing rs3745683(G>A), we unveiled the intricate molecular apparatus modulating *ANGPTL8* expression. Specifically, our investigations illuminated the role of the insulin-AKT1 signaling pathway in modulating *ANGPTL8* expression through the orchestration of nuclear translocation of the transcription factor YBX1. We unveiled a potential regulatory pathway underlying the dysregulation of lipid metabolism caused by OSA.

## Materials and Methods

### Clinical cohort

Individuals enrolled in this study were from SSHS cohort. The SSHS study adheres to the Declaration of Helsinki and is registered in the Chinese Clinical Trial Registry (ChiCTR1900025714). It has received ethical approval from the Ethics Committee of Shanghai Sixth People’s Hospital (Approval No.2021-KY-76), and the collection and preservation of genetic samples have been authorized by the China Human Genetic Resources Management Office (Approval No. 2022-BC0010).

The cohort consists of 5,438 OSA cases and 15,152 controls from Han Chinese. The individuals, genotyping and statistical analysis strategies were described in detail in our previous study [30]. Next, subjects from SSHS in this study, were screened according to the following exclusion criteria. (i) age less than 18 years old; (ii) history of OSA treatment; (iii) history of metabolic disorders treatment (i.e., hyperlipemia, T2MD); (iv) cardiovascular disease; (v) unavailable standard polysomnography data. A total 4,692 participants were ultimately analyzed in this study.

### Animal studies

All animal studies were supervised and approved by the Animal Care Committee of Shanghai Six People’s Hospital affiliated to Shanghai Jiao Tong University School of Medicine, and the animal welfare ethics acceptance number is No:2019-0237. Four-week-old C57BL/6N SPF male mice were purchased from GemPharmatech (China), The mice were randomly allocated to the experimental group or control group, and kept at humidity environment with free access to food and water in a standard light-dark cycle.

Before the adeno-associated virus (AAV) construction, the target sequence used against mouse *Ybx1* was screened from three candidates in the primary hepatocyte. AAV2/8 carrying mouse *Ybx1*-shRNA (AAV2/8-TBG-*Ybx1*-shRNA) and the control vectors (AAV2/8-TBG-*EV*) were constructed, amplified, and purified by Obio Technology (China). The target sequence used against mouse *Ybx1* was as follows: 5’-CCACGCAATTACCAGCAAA-3’. AAV diluted in saline and injected via the tail vein of mice (2 × 10^11^ vg per mouse). Three weeks later, immunoblotting or RT-qPCR were performed to examine the expression levels of *Ybx1* and *Angptl8* in mice livers. Then, the rest mice were fed with high fat-diet (HFD, 60% fat caloric content, Research Diets, D12492) and the body weights and food intakes were measured weekly. For 12 weeks of HFD feeding, we performed GTT on these mice. For 16 weeks of HFD feeding, mice were sacrificed under fasting conditions, plasm, adipose and liver of mice were collected for subsequent examination.

### Cellular and Molecular Studies

Genetic manipulation and metabolic measurement in human and mouse liver cell lines, as well as mouse adipocyte precursor cells, were used to investigate the role of the indicated genes in lipid metabolism. Luciferase reporter assays, CUT & Tag, DNA pull-down assays, and subsequent mass spectrometry were employed to study the transcriptional regulatory capacity of the indicated SNP and its underlying mechanisms. Quantitative PCR and Western blot analyses were performed to assess gene expression at the mRNA and protein levels. Detailed information on the materials and protocols used in the aforementioned studies is provided in the Supplementary Materials.

### Statistics

The clinical statistical analyses were performed using SPSS 26.0 software (IBM Corp., Armonk, NY, United States). The normally distributed data are presented as the means and standard deviation; skewed data are presented as the median (IQR), and categorical data are presented as the number (percentage). Differences in the baseline characteristics among different OSA severity degree groups were examined using one-way analysis of variance (ANOVA), non-parametric Kruskal–Wallis H test, or χ2 tests according to the type of data distribution. Baseline characteristics also tested by the polynomial linear trend test for continuous variables and the linear-by-linear association test for dichotomous variables.

For GWAS of OSA-related traits, the quantitative phenotypic variables were analyzed by linear regression under an additive genetic model after adjusting for age, sex, cigarette consumption, and alcohol consumption. Logistic regression analyses were performed to evaluate the relationship between polymorphisms and categorical variables.

For other statistical analysis, the results were presented as the mean ± SEM. Statistical significance among multiple groups was analyzed by one-way ANOVA, followed by Student’s t test using Prism 8 (GraphPad Software, Inc.) to compare the results between two groups. *P* < 0.05 was considered to be statistically significant.

## Results

### GWAS revealed novel genetic loci associated with serum lipid levels in OSA patients

To better elucidate the genetic factors underlying metabolic abnormalities in individuals with OSA, we performed a GWAS using SSHS cohort focusing on four serum lipid traits including total cholesterol (CHOL), triglyceride (TRIG), low-density lipoprotein cholesterol (LDL-C) and high-density lipoprotein cholesterol (HDL-C), and identified 18 genome-wide significant loci (*p* < 5 × 10^-8^) (Fig. 1a-1d).

**Figure 1.**
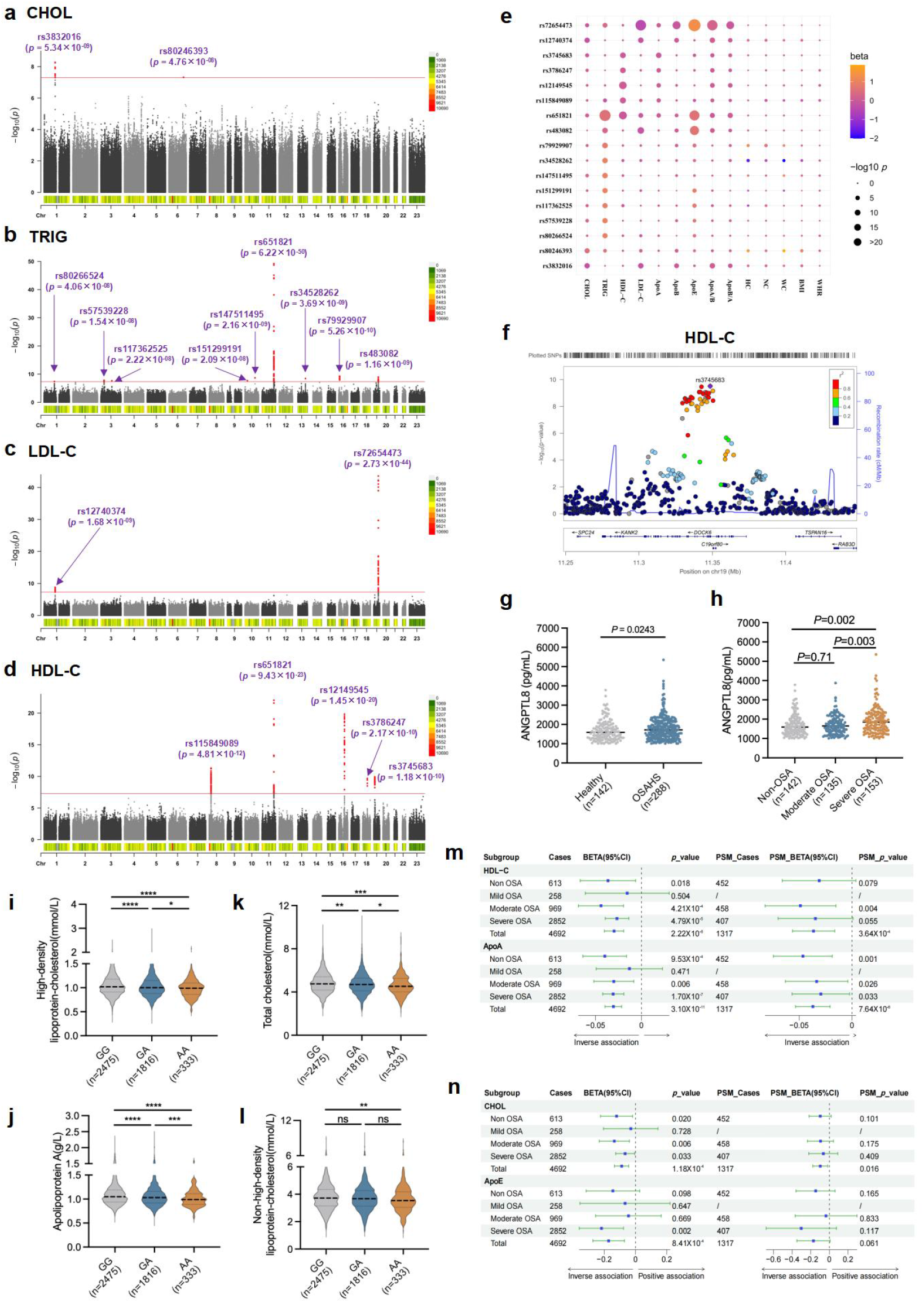
GWAS identified novel genetic loci associated with serum lipid traits in Han Chinese OSA patients. **(a-d)** Manhattan plot showing the genome-wide *p* values of association with serum CHOL **(a)**, TRIG **(b)**, LDL-C **(c)**, and HDL-C **(d)**. The red line shows the genome-wide significant threshold of *p* < 5 × 10^-8^. **(e)** Dotplot presents the associations between indicated variants and the quantitative traits. **(f)** Region plot of association between rs3745683 and HDL-C. The purple dot indicates rs3745683. **(g, h)** Serum ANGPTL8 levels in OSA patients. **(i-l)** The comparison of serum HDL-C **(i)**, CHOL **(k)**, ApoA **(j)**, and non HDL-C **(l)** levels among three genotypes of rs3745683. *p<0.05, **p<0.01, ***p<0.001, ****p<0.0001. **(m, n)** Effects of rs3745683(G>A) on serum HDL-C **(m)**, ApoA **(m)**, CHOL **(n)**, and ApoE **(n)** among different severity OSA patients. Beta and *p* value were obtained by linear regression analysis with or without adjusting for age, gender, cigarette consumption, and alcohol consumption.

Among them, 2 loci (rs3832016, and rs80246393) were identified for CHOL; 9 loci (rs80266524, rs57539228, rs117362525, rs151299191, rs147511495, rs651821, rs34528262, rs79929907, and rs483082) were identified for TRIG; 2 loci (rs12740374, and rs72654473) were identified for LDL-C; and the rest 5 loci (rs115849089, rs651821, rs12149545, rs3786247, and rs3745683) were identified for HDL-C (Table 1).

**Table 1.** Eighteen Genetic variant loci associated with serum CHOL, TRIG, LDL-C and HDL-C levels in GWAS cohort.

| Traits | SNP ID | Chr | A1 | A2 | $\beta$ | SE | *p* value | MAF |
| --- | --- | --- | --- | --- | --- | --- | --- | --- |
| CHOL | rs3832016 | 1 | C | CT | -0.2278 | 0.03897 | $5.34 \times 10^{-09}$ | 0.05824 |
| CHOL | rs80246393 | 6 | G | A | 0.5041 | 0.0922 | $4.76 \times 10^{-08}$ | 0.01101 |
| TRIG | rs80266524 | 1 | C | A | 0.9058 | 0.1648 | $4.06 \times 10^{-08}$ | 0.01118 |
| TRIG | rs57539228 | 3 | A | G | 0.9427 | 0.1664 | $1.54 \times 10^{-08}$ | 0.01082 |
| TRIG | rs117362525 | 3 | C | T | 0.8484 | 0.1514 | $2.22 \times 10^{-08}$ | 0.01429 |
| TRIG | rs151299191 | 10 | G | A | 0.9586 | 0.1708 | $2.09 \times 10^{-08}$ | 0.01019 |
| TRIG | rs147511495 | 10 | A | G | 0.9014 | 0.1504 | $2.16 \times 10^{-09}$ | 0.01373 |
| TRIG | rs651821 | 11 | C | T | 0.5633 | 0.03756 | $6.22 \times 10^{-50}$ | 0.2807 |
| TRIG | rs34528262 | 13 | T | G | 0.9393 | 0.159 | $3.69 \times 10^{-09}$ | 0.01197 |
| TRIG | rs79929907 | 16 | G | A | 0.779 | 0.1252 | $5.26 \times 10^{-10}$ | 0.01956 |
| TRIG | rs483082 | 19 | T | G | 0.2837 | 0.04654 | $1.16 \times 10^{-09}$ | 0.1889 |
| LDL-C | rs12740374 | 1 | T | G | -0.1934 | 0.03203 | $1.68 \times 10^{-09}$ | 0.05762 |
| LDL-C | rs72654473 | 19 | A | C | -0.3973 | 0.02821 | $2.73 \times 10^{-44}$ | 0.07703 |
| HDL-C | rs115849089 | 8 | A | G | 0.04752 | 0.006862 | $4.81 \times 10^{-12}$ | 0.1127 |
| HDL-C | rs651821 | 11 | C | T | -0.04699 | 0.004766 | $9.34 \times 10^{-23}$ | 0.2807 |
| HDL-C | rs12149545 | 16 | A | G | 0.05475 | 0.005868 | $1.45 \times 10^{-20}$ | 0.1616 |
| HDL-C | rs3786247 | 18 | G | T | 0.02829 | 0.004447 | $2.17 \times 10^{-10}$ | 0.4083 |
| HDL-C | rs3745683 | 19 | A | G | -0.0316 | 0.004896 | $1.18 \times 10^{-10}$ | 0.2695 |

A1, minor allele; A2, major allele; MAF, minor allele frequency.

Further analysis revealed no significant correlations between these loci and OSA respiratory events, blood oxygen saturation, or sleep architectures (Supplementary Fig. S1a - S1b). However, several loci demonstrated extensive associations with various metabolic traits. Notably, rs72654473 was closely associated with LDL-C, ApoB, and ApoE levels, while rs651821 showed strong associations with TRIG, HDL-C, and ApoE levels (Fig. 1e). The annotation and expression quantitative trait locus (eQTL) analysis of the 18 SNPs revealed that several loci or risk genes are widely characterized in several other GWAS of lipid traits, including *APOA5* (rs651821), *APOE* (rs72654473), *APOC1* (rs483082), *CELSR2* (rs3832016, rs12740374), *CETP* (rs12149545) and *LIPG* (rs3786247). Furthermore, eight SNPs, including the CHOL-associated SNP (rs80246393) and TRIG-associated SNPs (rs80266524, rs57539228, rs117362525, rs147511495, rs151299191, rs34528262, and rs79929907), along with their affected genes, have not been previously reported in relation to lipid metabolism (Supplementary Table S1).

The annotation analysis showed that rs3745683 is located in the upstream region of *ANGPTL8* gene (Fig. 1f). Clinical data reveal elevated levels of ANGPTL8 expression in the serum of OSA patients (Fig. 1g, 1h), and previous studies has also suggested that ANGPTL8 expression can serve as an indicator of OSA improvement following weight loss surgery [15], Consequently, we have selected rs3745683 for our subsequent investigations.

A subpopulation of 4,692 individuals from SSHS cohort, compromising 658 healthy controls, 258 Mild OSA, 969 Moderate OSA, 2,852 Severe OSA, were analyze to further validate the association of rs3745683 and serum lipid levels (Supplementary Table S2). The results revealed a significant association between the rs3745683 (G>A) variant and lower serum HDL-C, CHOL, ApoA and non HDL-C levels, which is a well characterized risk factor for CVD (Fig. 1i-1l, Supplementary Fig. S1g, S1h). Stratifying by OSA severity and adjusting for age, gender, cigarette consumption, and alcohol consumption, multiple linear regression analysis under an additive model revealed a significant inverse association (*p* < 0.05) between the rs3745683 and serum levels of HDL-C, CHOL, and ApoA across the three groups and the total population (Fig. 1m, 1n, Supplementary Table S3). Furthermore, compared to the non OSA group (β = -0.037, *p* = 0.018), rs3745683 exhibited a more pronounced regulatory effect on HDL-C levels in the moderate OSA group (β = -0.044, *p* = 4.21×10^-4^) (Fig. 1m, Supplementary Table S3). Propensity score matching analysis, which accounted for differences in sample size, confirmed that rs3745683 remained significantly associated with serum HDL-C levels in the moderate OSA group (*p* = 0.004) (Fig. 1m, Supplementary Tables S4, S5). In summary, data from clinical samples further validated the strong correlation between rs3745683 and lipid traits.

### Functional impact of SNP rs3745683 on transcriptional activity and its role in modulating lipid metabolism through ANGPTL8 expression

The locus annotation shows that rs3745683 is located 1,774 bp upstream of the transcription start site (TSS) of *ANGPTL8* and within the 14th intron of *DOCK6* (Fig. 2a). Analysis of Ensembl data revealed its localization within an active region associated with transcriptional regulation (Fig. 2a). Furthermore, eQTL analysis from GTEx Portal Database demonstrated a significant correlation between the expression of *DOCK6* and rs3745683 in various tissues, such as Adipose_Subcutaneous (β = 1.0, *p* = 1.3×10^-35^), Artery_Tibial (β = 1.2, *p* = 2.8×10^-54^), and Esophagus_Muscularis (β = 1.1, *p* = 7.8×10^-36^). Additionally, in Artery_Tibial (β = -0.29, *p* = 1.4×10^-3^) and adipose_naive (β = -0.56, *p* = 2.1×10^-3^), expression of *ANGPTL8* was also found to be associated based on Ensembl database (Supplementary Table S6). Therefore, we hypothesize that rs3745683 may influence the transcriptional activity of this chromosomal region. To address this hypothesis, we cloned DNA fragments of 1,200 bp containing the SNP into the 5’ end of a luciferase reporter gene (Fig. 2b, Supplementary Table S7). Luciferase activity was subsequently assessed in multiple lipid metabolism-related cell lines with reference allele of rs3745683, including human hepatocytes (HepG2, LO2), mouse hepatocytes (AML12), and mouse preadipocytes (Supplementary Fig. S2a, S2b). Remarkably, the results consistently demonstrated a significant increase in luciferase activity associated with the SNP-linked DNA fragments across different cell types. Furthermore, in comparison to the reference allele construct, the alternative construct significantly decreased transcriptional activity in all four cell lines (Fig. 2c-2f). These findings provide compelling evidence supporting the notion that rs3745683 may have a functional impact on transcriptional activity in relevant cellular contexts.

**Figure 2.**
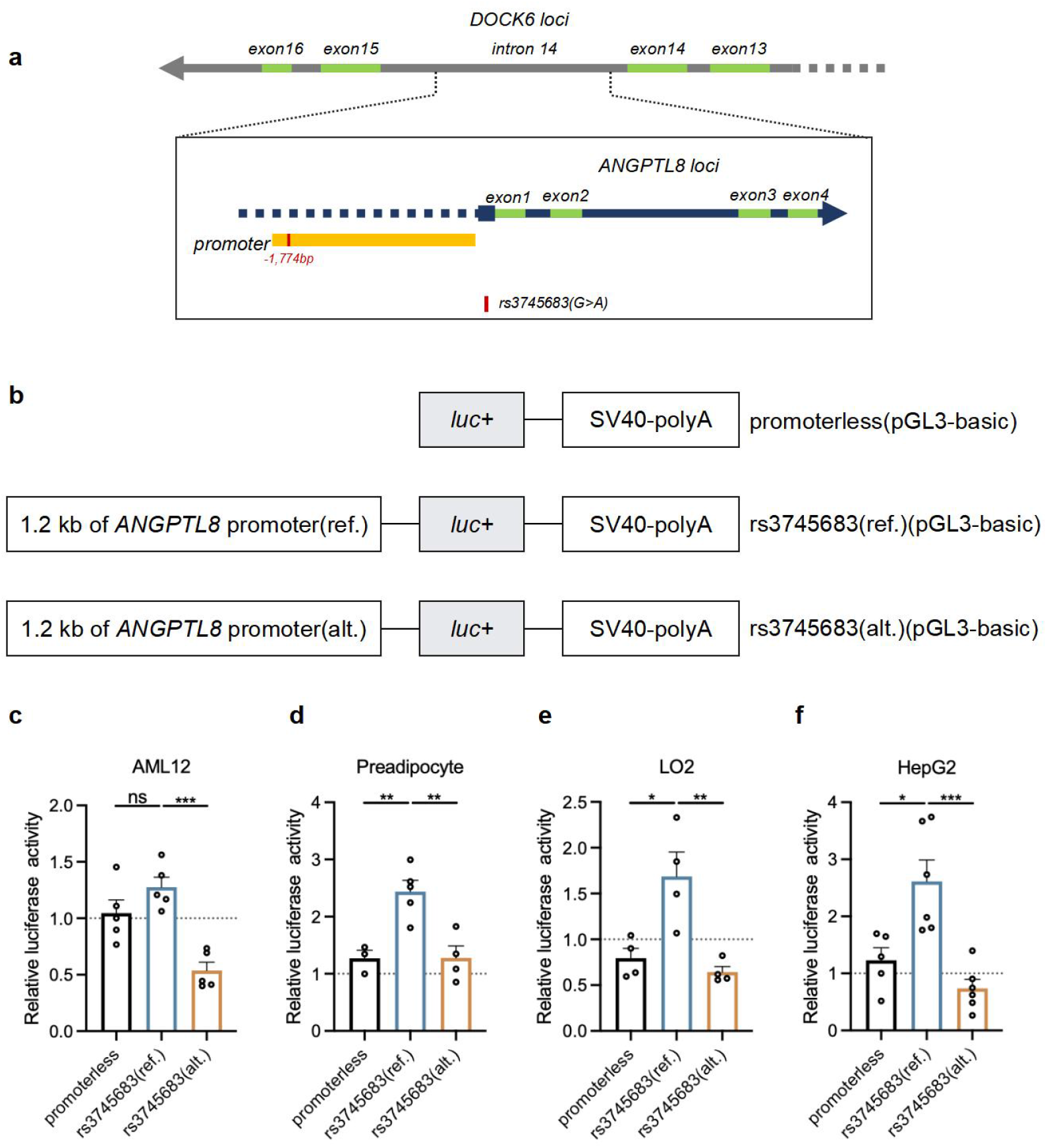
rs3745683 suppressed transcriptional activity of ANGPTL8 promoter. **(a)** Diagram of rs3745683 in ANGPTL8 promoter. **(b)** Scheme of vector constructs. **(c-f)** Mean (sem) transcriptional activity of *ANGPTL8* promoter with reference allele (G) or alternative allele (A) in AML12 **(c)**, mouse preadipocytes **(d)**, LO2 **(e)**, and HepG2 **(f)** cells. The y axis represents the relative luciferase activity.

Given the potential concurrent impact of rs3745683 on the expression of *DOCK6* and *ANGPTL8*, our investigation aimed to elucidate the key genes influenced by rs3745683 in the context of metabolic dysregulation. Previous studies have already established *ANGPTL8*, which highly expressed in liver and adipose tissue (Supplementary Fig. S3a), as a pivotal regulator of lipid metabolism. Our comparative analysis of ANGPTL8 expression in the serum of individuals carrying different alleles of rs3745683 revealed that homozygous individuals (n=31) carrying the alternative allele (A) exhibited significantly reduced levels of ANGPTL8 in their serum compared to both homozygous (n=39) and heterozygous individuals (n=56) with the reference allele (G) (Fig. 3a). Furthermore, CRISPR/Cas9 technology was used to conducted separate deletions of *Angptl8* in both mouse hepatocyte AML12 cells and adipocytes (Fig. 3b, 3c). Compared to wildtype control, the formation of lipid droplets and TRIG content were significantly decreased in *Angptl8*-sgRNAs transfected cells (Fig. 3d-3g), suggesting that *Angptl8* was required for triglyceride synthesis in both hepatocytes and adipocytes. Furthermore, we systematically excluded the involvement of *Dock6* deficiency in the observed TRIG synthesis changes (Supplementary Fig. S3b-S3d). As a consequence, we have substantiated that the SNP exerts its influence on lipid metabolism through the modulation of *ANGPTL8* expression.

**Figure 3.**
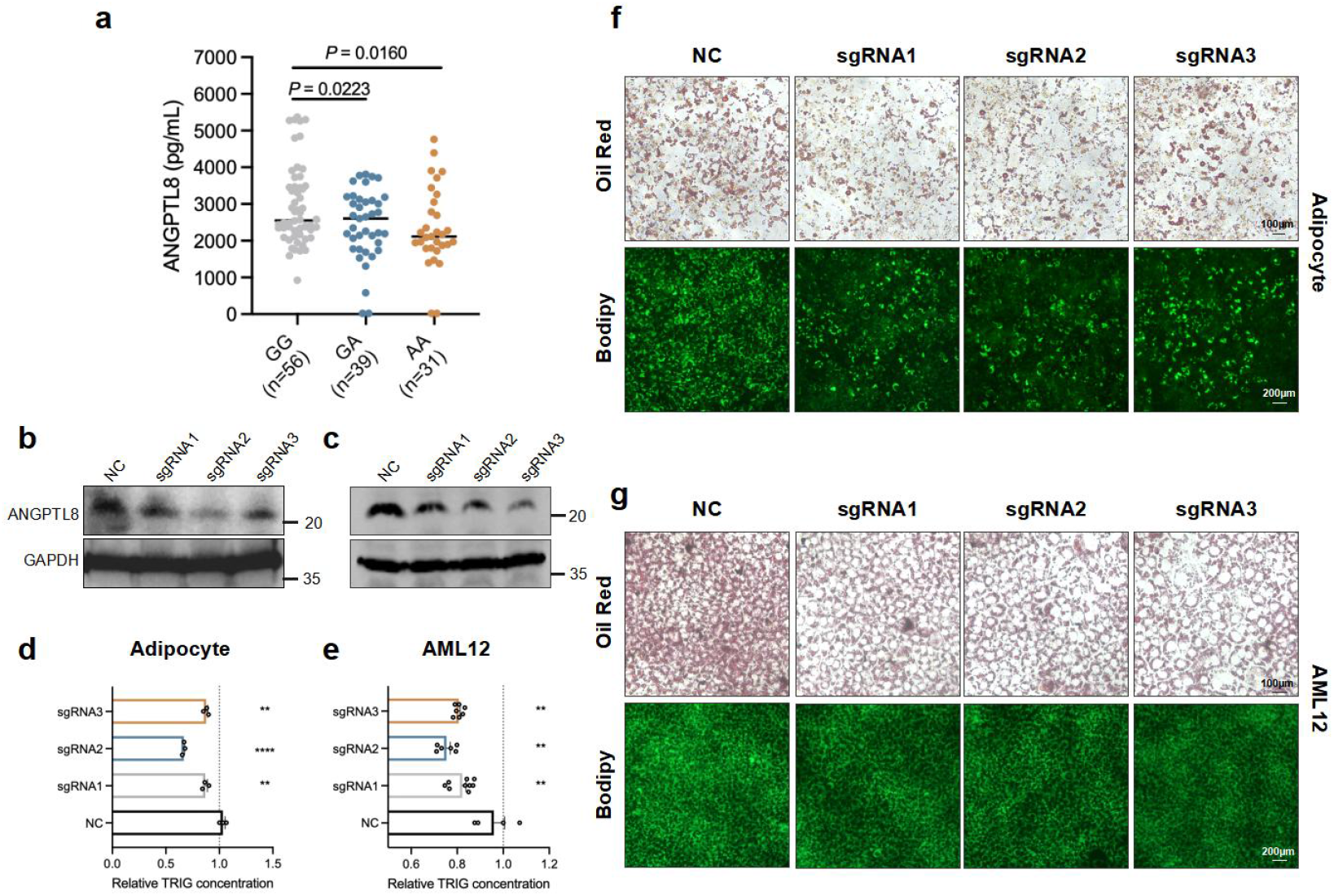
An essential role of ANGPTL8 in lipogenesis. **(a)** The comparison of serum ANGPTL8 level among three genotypes of rs3745683. **(b, c)** Protein levels of ANGPTL8 in *Angptl8-*sgRNA transfected adipocytes **(b)** and AML12 cells **(c)**. **(d)** Triglyceride content analysis of *Angptl8*-sgRNA transfected adipocytes differentiated for 6 days. **(e)** Triglyceride content analysis of *Angptl8*-sgRNA transfected AML12 cells differentiated for 1 day. **(f)** Oil red staining and Bodipy 493/503 dye staining of *Angptl8*-sgRNA transfected adipocytes differentiated for 6 days. **(g)** Oil red staining and Bodipy 493/503 dye staining of *Angptl8*-sgRNA transfected AML12 cells differentiated for 1 day.

### Identification of transcription factors YBX1, YBX2, PCBP1, and PCBP2 that bound to proximal sequence of rs3745683

To investigate whether rs3745683 influences the binding of adjacent DNA fragments to transcription factors or chromatin factors, biotin-labeled DNA probes, designed to mimic the DNA fragments associated with the SNP reference (G) and alternative (A) alleles, were synthesized at a length of 50 base pairs (bp) (Fig. 4a). Following incubation with HepG2 nuclear extracts, the resulting precipitates were subjected to streptavidin-based affinity chromatography. Subsequently, the protein bands specifically enriched in the precipitates containing the reference allele probe were subjected to mass spectrometry (MS) analysis. Through this method, we successfully identified the specific enrichment of transcription factors YBX1, YBX2, PCBP1, and PCBP2, collectively termed YYPP (Fig. 4b, 4c).

**Figure 4.**
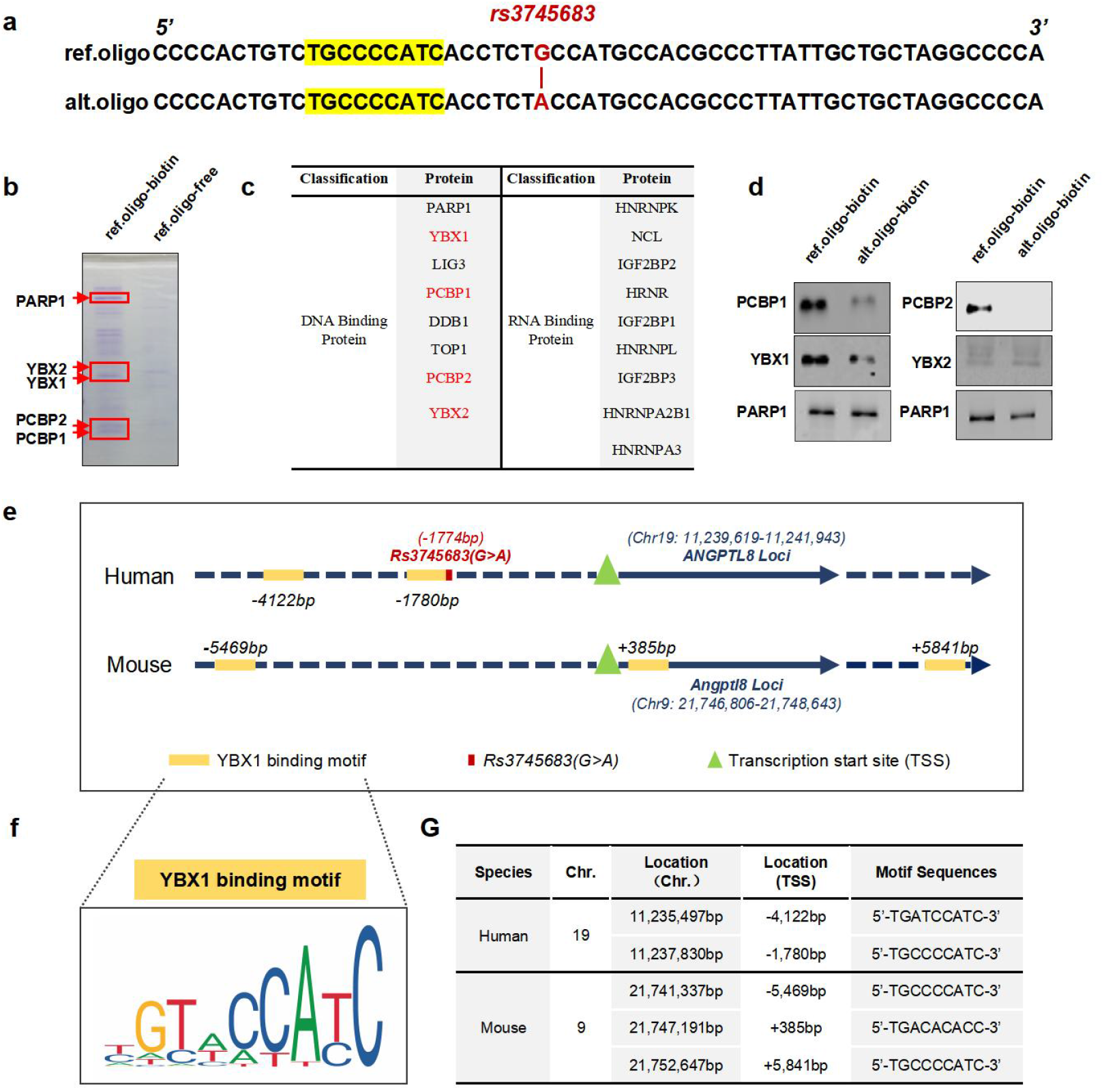
Identification of YBX1 as a transcriptional factor of rs3745683 proximal region. **(a)** DNA probe sequences containing rs3745683(G>A) locus. Highlight indicates the predicted YBX1 DNA binding motif. **(b)** Coomassie brilliant blue-stained acrylamide gel of a pulldown assay using oligonucleotides containing allele G of rs3745683. **(c)** Selected proteins in the ref.oligo-biotin precipitate identified by LC-MS analysis. **(d)** Immunoblots of a pulldown assay using oligonucleotides containing either allele G and A of rs3745683. **(e, g)** Predicted YBX1 DNA binding Motif in *ANGPTL8* gene among human and mouse. **(f)** Motif analysis of YBX1-bound sequence, available at https://jaspar.elixir.no/matrix/UN0139.1/.

We further validated the affinity precipitation results by conducting experiments in HepG2, which revealed a stronger enrichment of the transcription factor YBX1, PCBP1 and PCBP2 with the reference allele probe (Fig. 4d). Previous studies have indicated YYPP as a tightly bound protein complex [31–33], and YBX1 can directly bind to DNA sequences with a binding motif represented by (5’-TGTACCATC-3’) (Fig. 4f). Through sequence comparison, we identified that the proximal sequence to rs3745683 conforms to the binding motif characteristic of YBX1. Moreover, a further analysis of the upstream transcriptional regulatory sequence of mouse *Angptl8* revealed the presence of multiple YBX1 binding motifs (Fig. 4e-4g). Therefore, we hypothesize that YYPP are required for the transcriptional regulation of *ANGPTL8* expression cross species.

### YBX1 responds to insulin signaling and regulates *ANGPTL8* expression

Intermittent hypoxia and elevated insulin levels are two pathophysiological features in individuals with OSA, therefore, we investigated whether these factors affect the expression of *ANGPTL8* through YYPP [34]. Despite the ability of hypoxia stimulation to induce an increase in *ANGPTL8* expression (Supplementary Fig. S4k), the core transcription factor of hypoxia signaling, hypoxia inducible factor 1 subunit alpha (HIF1ɑ), was not enriched in the rs3745683 DNA fragment. Furthermore, Co-IP experiments did not reveal any interaction between HIF1ɑ and YYPP (Supplementary Fig. S4a-S4d). Additionally, luciferase reporter assays demonstrated that overexpression of YYPP did not alter the transcription of HIF1ɑ-responsive reporters (Supplementary Fig. S4e-S4h). Taken together, our results indicate that hypoxia signaling does not regulate *ANGPTL8* expression through the YYPP and rs3745683.

Since the little effect of HIF1ɑ on *ANGPTL8* expression by rs3745683, we focus on insulin signaling. Within our OSA cohort, we observed a substantial elevation in serum insulin levels and ANGPTL8 levels (Fig. 1g, 1h, Fig. 5a-5b). We conducted further analysis on a subpopulation of 123 individuals from the SSHS cohort to clarify the relationship between ANGPTL8 and insulin levels. Considerate the influence of insulin resistance, we then stratified HOMA-IR into quartiles corresponding to a HOMA-IR of Q1 (n = 30, HOMA-IR < 1.39), Q2 (n = 30, 1.39 ≤ HOMA-IR < 2.00), Q3 (n = 32, 2.00 ≤ HOMA-IR < 2.83), Q4 (n = 31, HOMA-IR ≥ 2.83). Linear regression analysis showed a significantly positive correlation (R^2^ = 0.1775, *p* = 0.0203) between serum ANGPTL8 and fasting insulin levels in group Q1 which performed a better insulin sensitivity compared to other groups. While, within the insulin resistance getting worse, the correlation between ANGPTL8 and insulin level getting weak (Fig. 5c). Collectively, these results implied that insulin facilitated ANGPTL8 expression and the decrease of sensitivity of insulin signaling pathway inhibited this progress.

**Figure 5.**
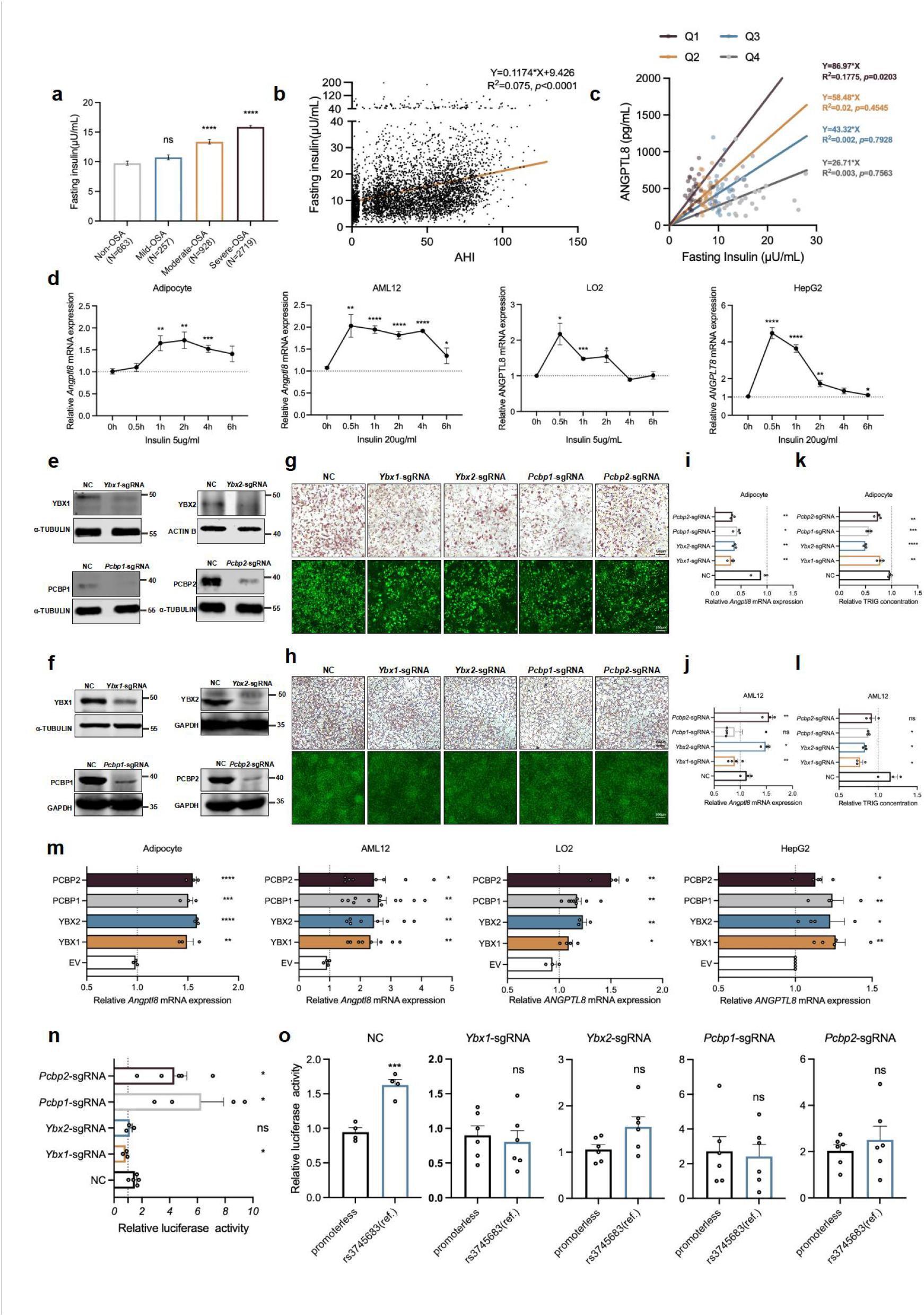
Effects of YYPP on Angptl8 expression and lipid metabolism. **(a-b)** Fasting insulin level in OSA patients **(a)** and correlation between insulin level and AHI **(b)**. **(c)** Linear association analysis of serum ANGPTL8 and fasting insulin level in different HOMA-IR group. **(d)** *ANGPTL8* mRNA levels in mouse adipocytes, AML12, LO2, and HepG2 cells following the stimulation of insulin over a 6-h period. **(e-f)** Immunoblots of YYPP-sgRNA knockout efficiency in mouse preadipocytes **(e)** and AML12 cells **(f)**. **(g-h)** Oil red staining and Bodipy 493/503 dye staining in YYPP-sgRNAs transfected adipocytes differentiated for 6 days **(g)**, and AML12 cells 200uM PA treated for 1 day **(h)**. **(i-j)** *Angptl8* mRNA levels in YYPP-sgRNAs transfected mouse adipocytes differentiated for 6 days **(i)**, and AML12 cells 200uM PA treated for 1 day **(j)**. **(k-l)** TRIG content in YYPP-sgRNAs transfected mouse adipocytes differentiated for 6 days **(k)**, and AML12 cells 200uM PA treated for 1 day **(l)**. **(m)** *ANGPTL8* mRNA levels in mouse adipocytes, AML12, LO2, and HepG2 cells after overexpression MYC-tagged YYPP for 2 days. **(n)** Mean (sem) transcriptional activity of *ANGPTL8* promoter with reference allele in YYPP deficiency preadipocytes. **(o)** Transcriptional activity comparison of vector constructs between promoterless and *ANGPTL8* promoter with reference allele in YYPP deficiency preadipocytes.

Additionally, both preadipocytes and hepatocytes demonstrated a significant upregulation of *ANGPTL8* expression in response to insulin signaling (Fig. 5d). To investigate whether the transcription factors YYPP mediate insulin regulation of *ANGPTL8*, we utilized CRISPR/Cas9 technology to perform targeted silence of YYPP expression in preadipocytes and AML12 cells (Fig. 5e, 5f). The results unequivocally showed that the depletion of YYPP in both cell types resulted in a substantial reduction in *ANGPTL8* expression and TRIG contents (Fig. 5g-5l). Consistently, overexpression of YYPP significantly increased *ANGPTL8* expression in all of four adipose and hepatic cell lines (Fig. 5m). Furthermore, luciferase reporter assays provided compelling additional evidence, supporting the notion that the loss of YYPP significantly impaired the transcriptional regulatory activity of *ANGPTL8* (Fig. 5n, 5o).

We then examined the cellular localization of YYPP in preadipocytes. In the untreated state, PCBP1 and PCBP2 were localized in the cell nucleus, whereas YBX1 and YBX2 were predominantly found in the cytoplasm (Fig. 6a). Upon insulin stimulation, YBX1 exhibited increased expression and translocated from the cytoplasm to the nucleus (Fig. 6a-6b). To investigate the signaling mechanisms involved in insulin stimulation, we examined the downstream activation of the PI3K-AKT and ERK MAP kinase pathways [35]. We used specific inhibitors, LY294002 and PD98059, to inhibit the PI3K-AKT and ERK MAP kinase pathways, respectively. Notably, inhibition of the PI3K-AKT pathway significantly reduced the nuclear translocation of YBX1, indicating that AKT kinase activity is crucial for nuclear YBX1 expression (Fig. 6c). This finding is supported by previous studies reporting AKT-mediated phosphorylation of YBX1 at Ser102 [36].

**Figure 6.**
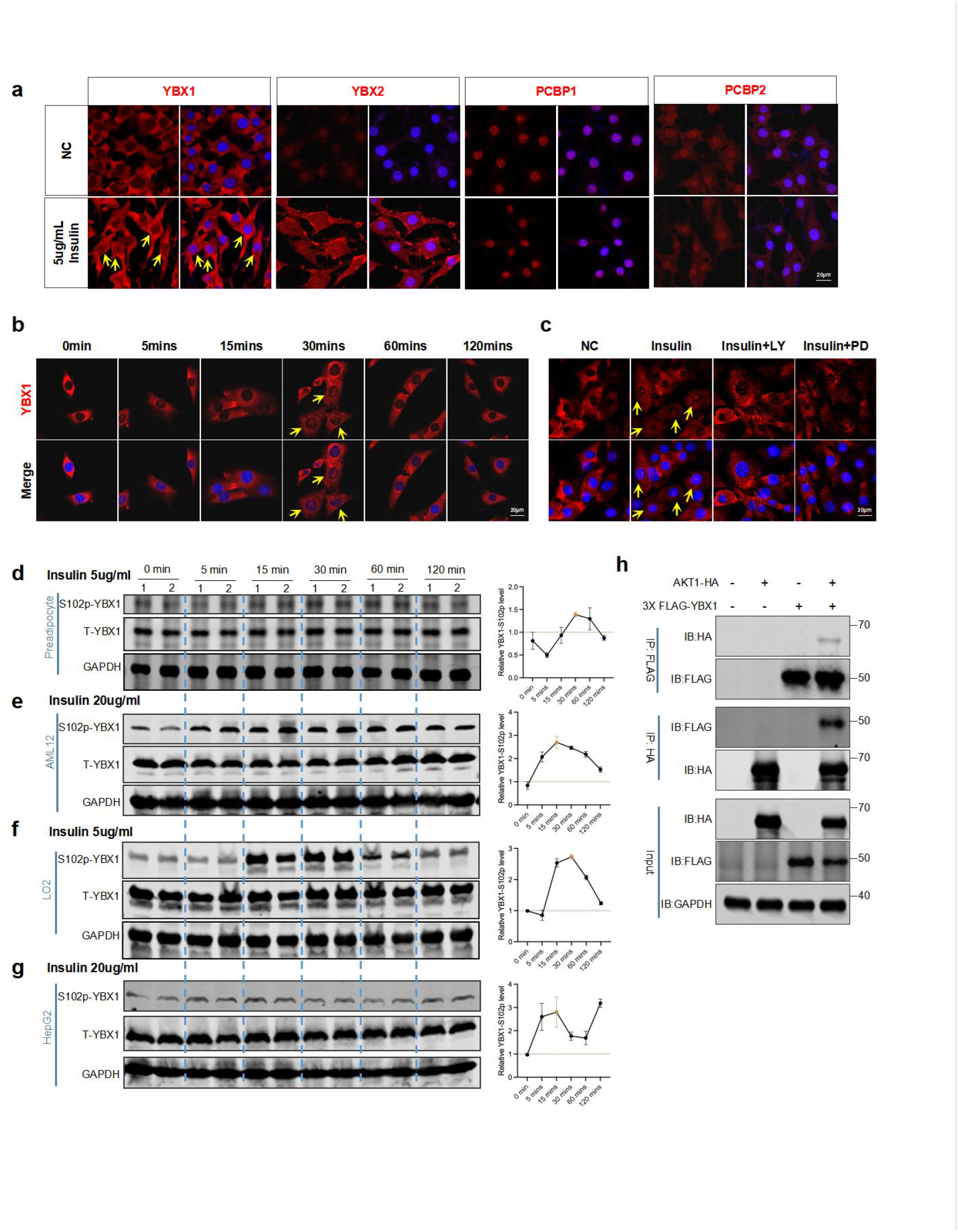
Insulin signaling triggers YBX1 phosphorylation and nuclear translocation through interaction between YBX1 and AKT. **(a)** Subcellular localization of YYPP in preadipocytes stimulated by 5µg/mL insulin for 24 h. **(b)** Subcellular localization of YBX1 in preadipocytes following the stimulation of 5µg/mL insulin over a 2-h period. **(c)** Subcellular localization of YBX1 in preadipocytes which stimulated by 5µg/mL insulin, 5µg/mL insulin+20µM LY294002, or 5µg/mL insulin+10µM PD98059 for 30mins, respectively. **(d-g)** YBX1-S102p levels in mouse preadipocytes **(d)**, AML12 **(e)**, LO2 **(f)**, and HepG2 **(g)** cells following the stimulation of insulin over a 2-h period. **(h)** Co-Immunoprecipitation analysis of 3×FLAG-YBX1 and AKT1-HA.

To validate this, we performed immunoblot experiments, which demonstrated clear insulin-induced phosphorylation of YBX1 at the Ser102 site across different cell types. Peak phosphorylation and subsequent nuclear translocation of YBX1 were observed 30 minutes after insulin treatment (Fig. 6b, 6d-6g). Furthermore, through immunoprecipitation experiments, we confirmed the protein interaction between AKT1 and YBX1, but did not observe interactions between YBX1 and AKT2 or AKT3 (Fig. 6h, Supplementary Fig. S4i, S4j). Collectively, these findings provide strong evidence that the insulin-AKT1 signaling pathway promotes the nuclear translocation of YBX1 through its phosphorylation.

To validate our observations, we generated two mutants of YBX1 at Ser102: the non-phosphorylatable Ser102Ala (S102A) and the phospho-mimicking Ser102Asp (S102D) mutants, along with mutants at other serine residues potentially involved in phosphorylation (Fig. 7a, Supplementary Fig. S5a). Intriguingly, the S102D and S176A mutants consistently exhibited nuclear localization even in the absence of insulin stimulation (Fig. 7a, Supplementary Fig. S5a). In *Ybx1*-sgRNA transfected cells, the S102D, S176A, and S176D mutants restored the expression of Angptl8 (Fig. 7b, 7c). In wildtype adipocytes and AML12 cells, transfection with the S102D, S176A, and S176D mutants significantly increased *Angptl8* expression and led to a notable elevation of TRIG content within the cells (Fig. 7d-7g).

**Figure 7.**
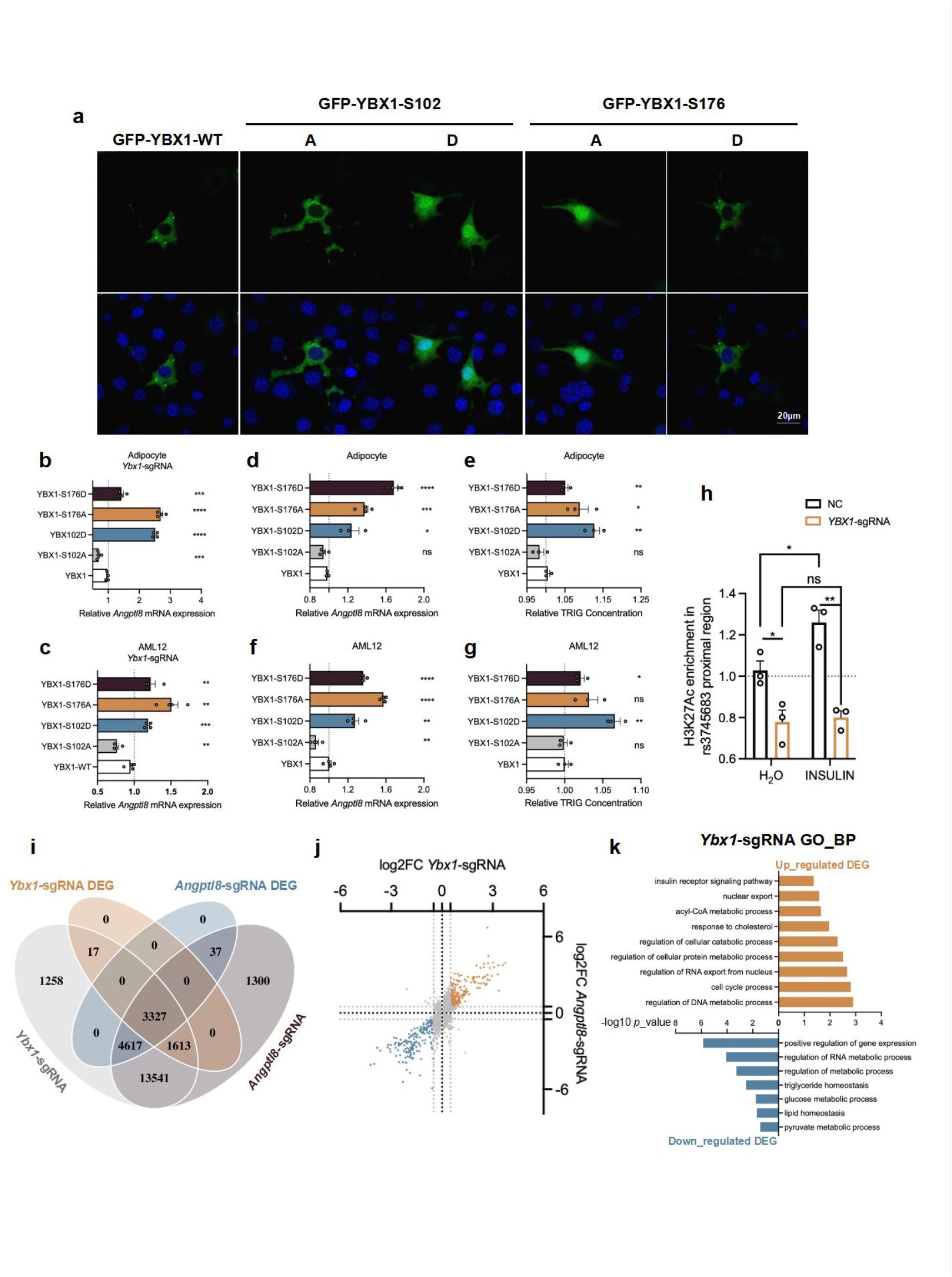
The phosphorylation dependent nuclear translocation of YBX1 promotes ANGPTL8 expression. **(a)** Subcellular localization of GFP-tagged YBX1 variations (WT, S102A, S102D, S176A, and S176D) in preadipocytes. The nuclei were stained with DAPI. These images were captured at 630 magnification. **(b-c)** *Angptl8* mRNA levels in *Ybx1*-sgRNA transfected mouse adipocytes **(b)**, and AML12 cells **(c)** which overexpressed 5 YBX1 variations respectively. **(d-e)** *Angptl8* mRNA levels **(d)**, and TRIG content **(e)** in mouse adipocytes which overexpressed 5 YBX1 variations respectively. **(f-g)** *Angptl8* mRNA levels **(f)** and TRIG content **(g)** in AML12 cells which overexpressed 5 YBX1 variations respectively. **(h)** With or without 20µg/mL insulin stimulation, H3K27Ac occupancy analysis of rs3745683 proximal region in HepG2 cells by CUT & Tag-qPCR. **(i)** Venn plot of gene expression profiles in *Ybx1*-sgRNA and *Angptl8*-sgRNA AML12 cells, DEG thresholds: *p* < 0.05. **(j)** DEGs (*p* < 0.05) in *Ybx1*-sgRNA and *Angptl8*-sgRNA AML12 cells. Gray line represented |log2FoldChange| = 0.5. **(k)** GO pathway enrichment analysis for biological process in *Ybx1*-sgRNA AML12 cells, up-regulated and down-regulated DEG thresholds: *p* < 0.05, |log2FoldChange| > 0.5.

Furthermore, we employed Cleavage Under Targets and Tagmentation (CUT & Tag) and real-time quantitative PCR (RT-qPCR) using an anti-H3K27Ac antibody to assess the transcriptional activity of the rs3745683 locus proximal region. Results showed that insulin increased the enrichment of the rs3745683 locus proximal region in HepG2 cells; however, this effect was inhibited by YBX1 deficiency (Fig. 7h). Taken together, these results indicate that YBX1 is essential for *Angptl8* expression, and the phosphorylation of YBX1 at Ser102 is critical for initiating this process.

We also performed RNA sequencing to analyze the gene expression profiles in *Ybx1*- and *Angptl8*-deficient AML12 cells (Supplementary Fig. S5b-S5c). Compared to the control group, 4,958 genes showed significant changes in expression (*p* < 0.05) in the *Ybx1*-deficient cells, and 7,982 genes were significantly differentially expressed in the *Angptl8*-deficient cells. Among these differentially expressed genes (DEGs), 1,389 genes were up-regulated in both groups (*p* < 0.05, log2FoldChange > 0), and 1,472 genes were down-regulated in both groups (*p* < 0.05, log2FoldChange < 0) (Fig. 7i-7j). Furthermore, functional enrichment analysis revealed that these DEGs were associated with lipid metabolism-related biological processes, such as “lipid homeostasis” and “cholesterol metabolic process” (Fig. 7k, Supplementary Fig. S5d). These results indicate similar gene expression profiles in *Ybx1*- and *Angptl8*-deficient hepatocytes, strongly suggesting that *Ybx1* affects lipid metabolism by regulating *Angptl8* expression.

### YBX1 regulates *Angptl8* expression and lipid metabolism in mouse liver

To further explore the effect on lipid metabolism of *Ybx1 in vivo*, we therefore performed the knock-down of *Ybx1* in liver by delivering AAV2/8 containing *Ybx1*-shRNA (AAV-*Ybx1-*shRNA) in C57BL/6J male mice, achieving approximately 80% specific deficiency in *Ybx1* levels in liver (Fig. 8a-8d). Consistently, we also observed about 20% and 50% decrease of *Angptl8* in mRNA level and protein level, respectively, in *Ybx1*-deficient mouse livers (Fig. 8b-8d).

**Figure 8.**
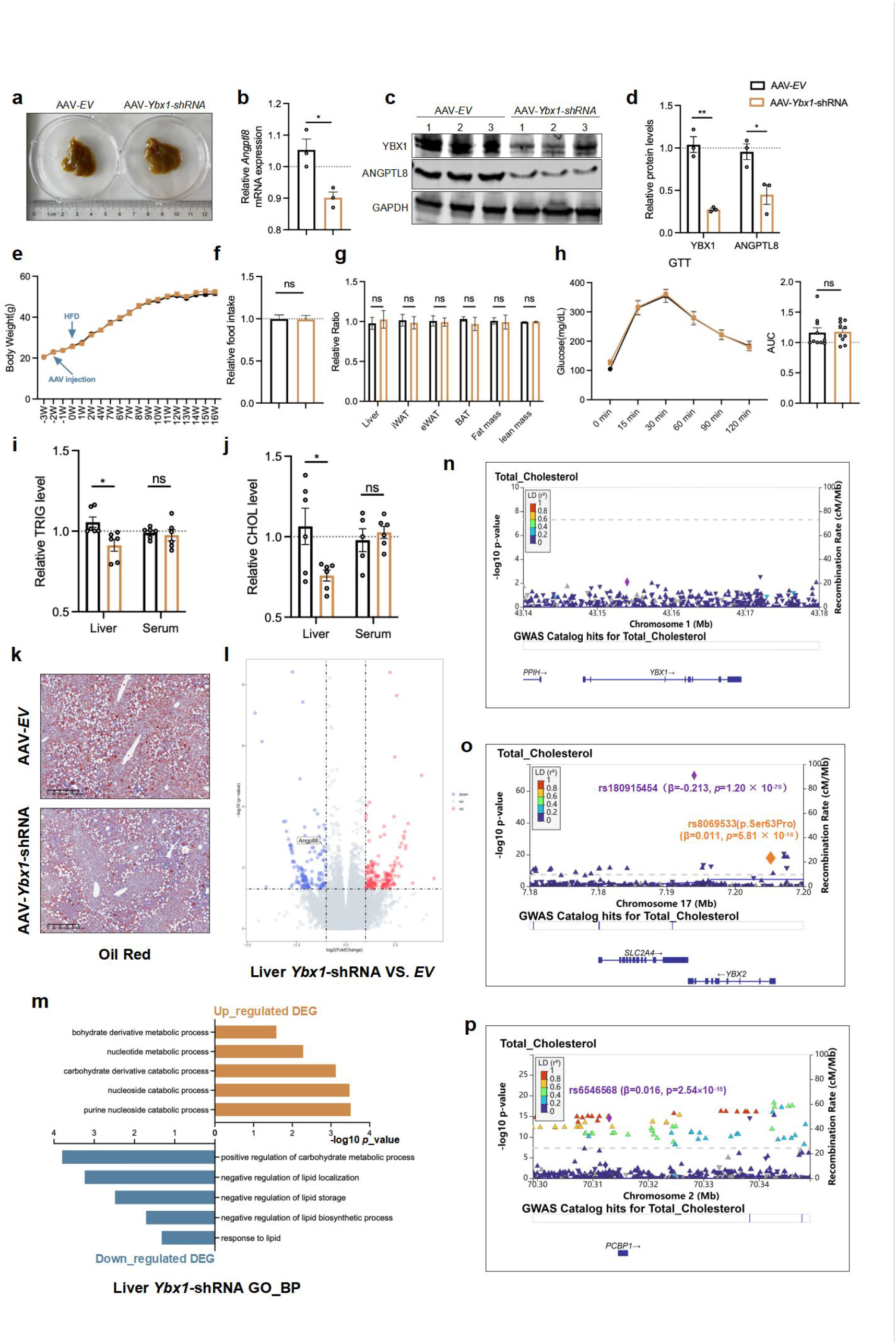
YBX1 regulates ANGPTL8 expression and liver lipid metabolism *in vivo*. **(a)** Pictures of AAV infected livers. **(b-d)** Levels of *Angptl8* mRNA **(b)** and protein **(c-d)** in AAV-*Ybx1*-shRNA mouse livers. **(e, f)** Body weights **(e)** and food intake **(f)** of AAV-*EV* and AAV-*Ybx1*-shRNA mice. **(g)** Relative tissue weight, fat mass, and lean mass of AAV-*EV* and AAV-*Ybx1*-shRNA mice under HFD for 16 weeks. **(h)** Glucose tolerance test of mice under HFD for 12 weeks**. (i-j)** TRIG **(i)** and CHOL **(j)** levels in serum and liver of AAV-*EV* and AAV-*Ybx1*-shRNA mice. **(k)** Oil red staining of mouse livers under HFD for 16 weeks. **(l)** Volcano plot showing the DEGs with statistical significance (thresholds: *p* < 0.05) and fold change (thresholds: |log2FoldChange| ≥ 1) in the livers of AAV-*EV* and AAV-*Ybx1*-shRNA mice. **(m)** GO pathway enrichment analysis for biological process in the livers of AAV-*Ybx1*-shRNA mice, up-regulated and down-regulated DEG thresholds: *p* < 0.05, |log2FoldChange| > 1. **(n-p)** Region plot of association between YBX1 locus **(n)**, YBX2 locus **(o)** and PCBP1 locus **(p)**, and CHOL trait.

We conducted a 16-week high-fat diet intervention on two genotypes of mice and examined their metabolic parameters. We found that the knockdown of *Ybx1* in the liver did not significantly affect the overall metabolic characteristics of the mice, including body weight, food intake, fat mass, lean mass, as well as the weights of metabolic organs including liver, inguinal white adipose tissue (iWAT), epididymal white adipose tissue (eWAT), and brown fat (Fig. 8e-8g). Additionally, under fasting conditions, there were no significant changes in serum TRIG and CHOL levels (Fig. 8i-8j). Meanwhile, mice of both genotypes after 12 weeks HDF treatment performed similarly in glucose tolerance test under fasting state (Fig. 8h). However, the levels of TRIG and CHOL were significantly decreased in *Ybx1*-deficient mouse liver, suggesting that *Ybx1* is required for the lipid synthesis in livers (Fig. 8i-8k). A further RNA-seq analysis showed the deficiency of YBX1 in mouse liver significantly decreased *Angptl8* mRNA level (log2Foldchange = -1.114, *p* = 0.002). The functional enrichment analysis showed that these significantly changed DEGs were associated with nucleotide or lipid metabolism-related biological processes, such as “nucleotide metabolic process” and “negative regulation of lipid biosynthetic process” (Fig. 8l-8m). These *in vivo* results showed that deficiency of *Ybx1* decreases ANGPTL8 expression in both mRNA and protein levels, and further improve TRIG and CHOL accumulation in liver.

To further investigate whether *YBX1* and its associated cofactors are involved in the regulation of human metabolism and the occurrence of metabolic diseases, we conducted an in-depth exploration using the EBI Metabolomics-related GWAS datasets. Unfortunately, our analysis did not identify any genetic variations in *YBX1* significantly associated with various metabolic traits (*p* < 5×10^-5^) (Fig. 8n). However, we did observe a genome-wide significant association between the non-coding SNP rs180915454 in *YBX2* and a decrease in total cholesterol (β = -0.213, *p* = 1.20×10^-70^) (Fig. 8o). Additionally, the missense variant rs8069533 (p. Ser63Pro) in *YBX2* showed a genome-wide significant association with an increase in total cholesterol (β = 0.011, *p* = 5.81×10^-18^) (Fig. 8o). Also, we identified an upstream SNP rs6546568 of *PCBP1* is a genome-wide significant variant of total cholesterol (β = 0.016, *p* = 2.54×10^-15^) (Fig. 8p). These genetic correlation data further suggest the significant involvement of *YBX1* and its associated factors in the regulation of human metabolism.

Collectively, our comprehensive findings shed light on the intricate molecular mechanisms involved in YBX1-mediated regulation of *ANGPTL8* expression.

## Discussion

A large number of studies, including meta-analyses have firmly established the profound impact of OSA, not only significantly elevating the risk of adverse health outcomes but also independently serving as a predictive factor for overall and cardiovascular mortality [37]. Additionally, individuals afflicted with OSA exhibit an enhanced susceptibility to the development of dyslipidemia and insulin resistance [38]. A twin study has underscored the substantial hereditary component contributing to the emergence of dyslipidemia in OSA patients [39]. Recent genetic investigations conducted within OSA populations have shed light on the role played by genetic variations, including those within the leptin receptor and ApoA5, in the manifestation of dyslipidemia among OSA patients [40, 41].

Nevertheless, the precise mechanisms by which OSA instigates disruptions in lipid metabolism have remained largely uncharted. Our study has brought to the forefront a noteworthy correlation between genetic variations in the endocrine hormone ANGPTL8 and lipid irregularities in OSA patients. Moreover, we have observed a significant elevation in ANGPTL8 levels in OSA patients, with both of OSA’s pathophysiological hallmarks, intermittent hypoxia and insulin resistance, contributing to an upregulation in ANGPTL8 expression. We have further elucidated how the insulin-AKT1-YBX1 pathway, by fostering the expression of ANGPTL8, subsequently amplifies lipid synthesis in the liver and adipose tissues. Our findings serve to enhance our comprehension of the molecular mechanisms through which OSA induces abnormalities in lipid metabolism.

Our research, conducted at both the human genetic and molecular cellular levels, unambiguously supports the pivotal role of ANGPTL8, a serum hormone, in the context of dyslipidemia in OSA. This observation aligns with the recent body of research that underscores the extensive yet highly conserved role of ANGPTL8 in metabolic regulation. ANGPTL8, a serum hormone secreted by both the liver and adipose tissues, further modulates the glycemic and lipidic processes in muscle, liver, and adipose tissues through endocrine mechanisms. Notably, recent studies by Siyu chen et al. have revealed that ANGPTL8 can interact with hepatic surface receptor PirB, thereby influencing the expression of circadian genes in hepatocytes, with implications for hepatic metabolism [42]. Moreover, multiple investigations have underscored the pivotal role of ANGPTL8 in the onset and progression of various metabolic disorders. For instance, Zongli Zhang and colleagues unveiled that ANGPTL8 exerts a promotive effect on the development of non-alcoholic fatty liver disease [16]. Targeted strategies aimed at inhibiting ANGPTL8 have already demonstrated promising effects in models of dyslipidemia in humanized mice [16]. Collectively, our research also suggests that the targeted inhibition of ANGPTL8 may hold potential therapeutic value in addressing metabolic syndrome.

Y-box binding protein 1 (YBX1), a member of the DNA- and RNA-binding protein family, has garnered significant attention due to its involvement in malignant cell transformation, tumor aggressiveness, and its potential as a therapeutic target in cancer and inflammation [43]. Initially recognized as a DNA transcription factor for its binding to a DNA nucleotide sequence known as the Y-box (5’-CTGATTGGC/TC/TAA-3’) located in gene promoters [44], YBX1 has since been attributed to a range of functions, including DNA repair, pre-mRNA splicing, translation, and packaging [45]. Recent studies have unveiled YBX1’s ability to influence gene expression both within and outside the context of the Y-box sequence. Notably, DNA pulldown and LC‒MS analyses conducted in our study have indicated that YBX1, in concert with YBX2, PCBP1, and PCBP2, represents potential transcription factors binding to the rs3745683 region. Furthermore, the binding affinities of YBX1, PCBP1, and PCBP2 to rs3745683 (G>A) are diminished in the presence of this genetic variation.

Emerging research has illuminated YBX1’s role not only in tumorigenesis but also in the regulation of thermogenic gene expression in adipocytes and the transformation of white adipose tissue into brown adipose tissue [46, 47]. Another study has demonstrated that YBX1 promotes brown adipogenesis and thermogenic activity through PINK1/PRKN-mediated mitophagy [48]. These findings underscore the multifaceted functions of YBX1 in adipose tissue biology. Our study, in which we employed CRISPR/Cas9 to generate TFs-deficiency adipocyte and hepatocyte cell lines, revealed that the depletion of YBX1 significantly reduced *Angptl8* expression, intracellular lipid droplets, and triglyceride content. Furthermore, our prior research has shown that serum fasting insulin levels are positively correlated with the severity of OSA [49]. Thus, it is reasonable to infer that YBX1, a responsive factor to insulin signaling, exerts regulatory control over *ANGPTL8* expression by binding to the rs3745683 region, thereby influencing adipocyte lipid metabolism in OSA. Nevertheless, it is important to note that our study is grounded mostly *in vitro* experiments, and further *in vivo* investigations are imperative for a comprehensive understanding of these mechanisms.

### CRediT authorship contribution statement

Yuenan Liu: Methodology, Investigation, Formal analysis, Data analysis, Writing-original draft. Haolin Yuan: Investigation, Validation. Anzhao Wang: Investigation, Validation. Shengming Wang: Data analysis. Xu Xu: Funding acquisition, Methodology, Investigation. Junhui Hu: Investigation. Jinhong Shen: Funding acquisition, Methodology, Investigation. Yiming Hu: Funding acquisition, Methodology, Investigation. Xinyi Li: Funding acquisition, Investigation. Niannian Li: Methodology, Data analysis. Zhenfei Gao: Methodology, Investigation. Xiaoxu Zhang: Investigation. Xiaoman Zhang: Investigation. Huajun Xu: Methodology. Hongliang Yi: Funding acquisition, Methodology. Jian Guan: Funding acquisition, Supervision. Zhiqiang Li: Methodology, Supervision. Yongxu Zhao: Funding acquisition, Methodology, Investigation, Supervision. Shankai Yin: Funding acquisition, Supervision, Methodology, Writing-original draft. Feng Liu: Funding acquisition, Methodology, Supervision, Writing-original draft.

## Declaration of competing interest

All authors in this study declare no competing interests.

### Declaration of generative AI and AI-assisted technologies in the writing process

During the preparation of this work the authors used OpenAI’s tool ChatGPT in order to improve the English grammar. After using this tool, the authors reviewed and edited the content as needed and take full responsibility for the content of the publication.

## Data Availability Statement

Full summary statistics for the gwas study of SSHS can be accessed from https://figshare.com/ (DOI: 10.6084/m9.figshare.20033246) and Bio-X institutes website (http://analysis.bio-x.cn/gwas/). The raw genetic data for each individual from our analyses could also be accessed under the Data Access Agreement (https://figshare.com/, DOI: 10.6084/m9.figshare.20033243 and http://galaxy.bio-x.cn/library/list#folders/Ff2db41e1fa331b3e). Additional raw data can be obtained upon request from the authors.

## Acknowledgements

We would like to thank Professor Junli Liu from Shanghai Diabetes Institute, Department of Endocrinology and Metabolism, Shanghai Jiao Tong University Affiliated Sixth People’s Hospital for his assistance in indirect experiment. This study was also supported by grants-in-aid from Ministry of Science and Technology of the People’ s Republic of China (2021ZD0201900), National Natural Science Foundation of China (81971240, 82070824, 82171125, 81970869, 81970870, 82271153, 82000967, 82101205, 82100105, 82301291, 82300962), Shanghai Municipal Commission of Science and Technology (Grant No.18DZ2260200), the Interdisciplinary Program of Shanghai Jiao Tong University (YG2023LC11), Youth Innovation Promotion Association of CAS (2023289), Shanghai Sailing Program grant (23YF1431900). The funding sources had no role in design, conduct, analysed, and interpreted of this study.

## References

[1] Åberg, F., et al., Alcohol consumption and metabolic syndrome: Clinical and epidemiological impact on liver disease. J Hepatol, 2023. 78(1): p. 191–206. DOI: 10.1016/j.jhep.2022.08.030.

[2] Mesarwi, O.A., R. Loomba, and A. Malhotra, Obstructive Sleep Apnea, Hypoxia, and Nonalcoholic Fatty Liver Disease. Am J Respir Crit Care Med, 2019. 199(7): p. 830–841. DOI: 10.1164/rccm.201806-1109TR.

[3] Gaines, J., et al., Obstructive sleep apnea and the metabolic syndrome: The road to clinically-meaningful phenotyping, improved prognosis, and personalized treatment. Sleep Med Rev, 2018. 42: p. 211–219. DOI: 10.1016/j.smrv.2018.08.009.

[4] Xu, S., et al., The association between obstructive sleep apnea and metabolic syndrome: a systematic review and meta-analysis. BMC Pulm Med, 2015. 15: p. 105. DOI: 10.1186/s12890-015-0102-3.

[5] Trzepizur, W., et al., Sleep Apnea-Specific Hypoxic Burden, Symptom Subtypes, and Risk of Cardiovascular Events and All-Cause Mortality. Am J Respir Crit Care Med, 2022. 205(1): p. 108–117. DOI: 10.1164/rccm.202105-1274OC.

[6] Guo, C., et al., ANGPTL8 in metabolic homeostasis: more friend than foe? Open Biol, 2021. 11(9): p. 210106. DOI: 10.1098/rsob.210106.

[7] Zhang, R. and A.B. Abou-Samra, Emerging roles of Lipasin as a critical lipid regulator. Biochem Biophys Res Commun, 2013. 432(3): p. 401–5. DOI: 10.1016/j.bbrc.2013.01.129.

[8] Meng, M., et al., Zinc finger protein ZNF638 regulates triglyceride metabolism via ANGPTL8 in an estrogen dependent manner. Metabolism, 2024. 152: p. 155784. DOI: 10.1016/j.metabol.2024.155784.

[9] Guo, T., et al., Association of the angiopoietin-like protein 8 rs2278426 polymorphism and several environmental factors with serum lipid levels. Mol Med Rep, 2015. 12(3): p. 3285–3296. DOI: 10.3892/mmr.2015.3825.

[10] Crujeiras, A.B., et al., Interplay of atherogenic factors, protein intake and betatrophin levels in obese-metabolic syndrome patients treated with hypocaloric diets. Int J Obes (Lond), 2016. 40(3): p. 403–10. DOI: 10.1038/ijo.2015.206.

[11] Luo, M. and D. Peng, ANGPTL8: An Important Regulator in Metabolic Disorders. Front Endocrinol (Lausanne), 2018. 9: p. 169. DOI: 10.3389/fendo.2018.00169.

[12] Wang, D., et al., Angiopoietin-Like Protein 8/Leptin Crosstalk Influences Cardiac Mass in Youths With Cardiometabolic Risk: The BCAMS Study. Front Endocrinol (Lausanne), 2021. 12: p. 788549. DOI: 10.3389/fendo.2021.788549.

[13] Chen, X., et al., Circulating betatrophin levels are increased in patients with type 2 diabetes and associated with insulin resistance. J Clin Endocrinol Metab, 2015. 100(1): p. E96–100. DOI: 10.1210/jc.2014-2300.

[14] Al-Terki, A., et al., Increased Level of Angiopoietin Like Proteins 4 and 8 in People With Sleep Apnea. Front Endocrinol (Lausanne), 2018. 9: p. 651. DOI: 10.3389/fendo.2018.00651.

[15] Song, Z., et al., Decreased serum betatrophin may correlate with the improvement of obstructive sleep apnea after Roux-en-Y Gastric Bypass surgery. Sci Rep, 2021. 11(1): p. 1808. DOI: 10.1038/s41598-021-81379-1.

[16] Zhang, Z., et al., ANGPTL8 accelerates liver fibrosis mediated by HFD-induced inflammatory activity via LILRB2/ERK signaling pathways. J Adv Res, 2023. 47: p. 41–56. DOI: 10.1016/j.jare.2022.08.006.

[17] Lee, Y.H., et al., Association between betatrophin/ANGPTL8 and non-alcoholic fatty liver disease: animal and human studies. Sci Rep, 2016. 6: p. 24013. DOI: 10.1038/srep24013.

[18] Chen, J., et al., In vivo targeted delivery of ANGPTL8 gene for beta cell regeneration in rats. Diabetologia, 2015. 58(5): p. 1036–44. DOI: 10.1007/s00125-015-3521-z.

[19] DiDonna, N.M., Y.Q. Chen, and R.J. Konrad, Angiopoietin-like proteins and postprandial partitioning of fatty acids. Curr Opin Lipidol, 2022. 33(1): p. 39–46. DOI: 10.1097/MOL.0000000000000798.

[20] Zhang, R. and K. Zhang, An updated ANGPTL3-4-8 model as a mechanism of triglyceride partitioning between fat and oxidative tissues. Prog Lipid Res, 2022. 85: p. 101140. DOI: 10.1016/j.plipres.2021.101140.

[21] Henneman, P., et al., Genetic architecture of plasma adiponectin overlaps with the genetics of metabolic syndrome-related traits. Diabetes Care, 2010. 33(4): p. 908–13. DOI: 10.2337/dc09-1385.

[22] Fall, T. and E. Ingelsson, Genome-wide association studies of obesity and metabolic syndrome. Mol Cell Endocrinol, 2014. 382(1): p. 740–757. DOI: 10.1016/j.mce.2012.08.018.

[23] Aguilera, C.M., J. Olza, and A. Gil, Genetic susceptibility to obesity and metabolic syndrome in childhood. Nutr Hosp, 2013. 28 Suppl 5: p. 44–55. DOI: 10.3305/nh.2013.28.sup5.6917.

[24] Norris, J.M. and S.S. Rich, Genetics of glucose homeostasis: implications for insulin resistance and metabolic syndrome. Arterioscler Thromb Vasc Biol, 2012. 32(9): p. 2091–6. DOI: 10.1161/ATVBAHA.112.255463.

[25] Su, X., Y. Cheng, and B. Wang, ANGPTL8 in cardio-metabolic diseases. Clin Chim Acta, 2021. 519: p. 260–266. DOI: 10.1016/j.cca.2021.05.017.

[26] Cannon, M.E., et al., Trans-ancestry Fine Mapping and Molecular Assays Identify Regulatory Variants at the ANGPTL8 HDL-C GWAS Locus. G3 (Bethesda), 2017. 7(9): p. 3217–3227. DOI: 10.1534/g3.117.300088.

[27] Hanson, R.L., et al., The Arg59Trp variant in ANGPTL8 (betatrophin) is associated with total and HDL-cholesterol in American Indians and Mexican Americans and differentially affects cleavage of ANGPTL3. Mol Genet Metab, 2016. 118(2): p. 128–37. DOI: 10.1016/j.ymgme.2016.04.007.

[28] Lee, Y.H., et al., APOE and KLF14 genetic variants are sex-specific for low high-density lipoprotein cholesterol identified by a genome-wide association study. Genet Mol Biol, 2022. 45(1): p. e20210280. DOI: 10.1590/1678-4685-GMB-2021-0280.

[29] Quagliarini, F., et al., Atypical angiopoietin-like protein that regulates ANGPTL3. Proc Natl Acad Sci U S A, 2012. 109(48): p. 19751–6. DOI: 10.1073/pnas.1217552109.

[30] Xu, H., et al., Genome-Wide Association Study of Obstructive Sleep Apnea and Objective Sleep-related Traits Identifies Novel Risk Loci in Han Chinese Individuals. Am J Respir Crit Care Med, 2022. 206(12): p. 1534–1545. DOI: 10.1164/rccm.202109-2044OC.

[31] Funke, B., et al., The mouse poly(C)-binding protein exists in multiple isoforms and interacts with several RNA-binding proteins. Nucleic Acids Res, 1996. 24(19): p. 3821–8. DOI: 10.1093/nar/24.19.3821.

[32] Rosenbluh, J., et al., Genetic and Proteomic Interrogation of Lower Confidence Candidate Genes Reveals Signaling Networks in β-Catenin-Active Cancers. Cell Syst, 2016. 3(3): p. 302–316.e4. DOI: 10.1016/j.cels.2016.09.001.

[33] Huttlin, E.L., et al., Dual proteome-scale networks reveal cell-specific remodeling of the human interactome. Cell, 2021. 184(11): p. 3022–3040.e28. DOI: 10.1016/j.cell.2021.04.011.

[34] Dempsey, J.A., et al., Pathophysiology of sleep apnea. Physiol Rev, 2010. 90(1): p. 47–112. DOI: 10.1152/physrev.00043.2008.

[35] Siddle, K., Signalling by insulin and IGF receptors: supporting acts and new players. J Mol Endocrinol, 2011. 47(1): p. R1–10. DOI: 10.1530/JME-11-0022.

[36] Alkrekshi, A., et al., A comprehensive review of the functions of YB-1 in cancer stemness, metastasis and drug resistance. Cell Signal, 2021. 85: p. 110073. DOI: 10.1016/j.cellsig.2021.110073.

[37] Fu, Y., et al., Meta-analysis of all-cause and cardiovascular mortality in obstructive sleep apnea with or without continuous positive airway pressure treatment. Sleep Breath, 2017. 21(1): p. 181–189. DOI: 10.1007/s11325-016-1393-1.

[38] Framnes, S.N. and D.M. Arble, The Bidirectional Relationship Between Obstructive Sleep Apnea and Metabolic Disease. Front Endocrinol (Lausanne), 2018. 9: p. 440. DOI: 10.3389/fendo.2018.00440.

[39] Meszaros, M., et al., Obstructive sleep apnea and hypertriglyceridaemia share common genetic background: Results of a twin study. J Sleep Res, 2020. 29(4): p. e12979. DOI: 10.1111/jsr.12979.

[40] Popko, K., et al., Frequency of distribution of leptin receptor gene polymorphism in obstructive sleep apnea patients. J Physiol Pharmacol, 2007. 58 Suppl 5(Pt 2): p. 551–61.

[41] Bielicki, P., et al., Impact of polymorphism of selected genes on the diagnosis of type 2 diabetes in patients with obstructive sleep apnea. Pol Arch Intern Med, 2019. 129(1): p. 6–11. DOI: 10.20452/pamw.4406.

[42] Chen, S., et al., Angptl8 mediates food-driven resetting of hepatic circadian clock in mice. Nat Commun, 2019. 10(1): p. 3518. DOI: 10.1038/s41467-019-11513-1.

[43] Lyabin, D.N., I.A. Eliseeva, and L.P. Ovchinnikov, YB-1 protein: functions and regulation. Wiley Interdiscip Rev RNA, 2014. 5(1): p. 95–110. DOI: 10.1002/wrna.1200.

[44] Wolffe, A.P., Structural and functional properties of the evolutionarily ancient Y-box family of nucleic acid binding proteins. Bioessays, 1994. 16(4): p. 245–51. DOI: 10.1002/bies.950160407.

[45] Ceman, S., R. Nelson, and S.T. Warren, Identification of mouse YB1/p50 as a component of the FMRP-associated mRNP particle. Biochem Biophys Res Commun, 2000. 279(3): p. 904–8. DOI: 10.1006/bbrc.2000.4035.

[46] Rabiee, A., et al., White adipose remodeling during browning in mice involves YBX1 to drive thermogenic commitment. Mol Metab, 2021. 44: p. 101137. DOI: 10.1016/j.molmet.2020.101137.

[47] Park, J.H., et al., A multifunctional protein, EWS, is essential for early brown fat lineage determination. Dev Cell, 2013. 26(4): p. 393–404. DOI: 10.1016/j.devcel.2013.07.002.

[48] Wu, R., et al., RNA-binding protein YBX1 promotes brown adipogenesis and thermogenesis via PINK1/PRKN-mediated mitophagy. Faseb j, 2022. 36(3): p. e22219. DOI: 10.1096/fj.202101810RR.

[49] Zou, J., et al., Independent relationships between cardinal features of obstructive sleep apnea and glycometabolism: a cross-sectional study. Metabolism, 2018. 85: p. 340–347. DOI: 10.1016/j.metabol.2017.11.021.

